# Potential of ChatGPT in Youth Mental Health Emergency Triage: Comparative Analysis with Clinicians

**DOI:** 10.1101/2025.01.06.24319771

**Authors:** Samanvith Thotapalli, Musa Yilanli, Ian McKay, William Leever, Eric Youngstrom, Karah Harvey-Nuckles, Kimberly Lowder, Steffanie Schweitzer, Erin Sunderland, Daniel I. Jackson, Emre Sezgin

## Abstract

**Background:** Large language models (LLMs), such as GPT-4, are increasingly integrated into healthcare to support clinicians in making informed decisions. Given ChatGPT’s potential, it is necessary to explore such applications as a support tool, particularly within mental health telephone triage services. This study evaluates whether GPT-4 models can accurately triage psychiatric emergency vignettes and compares its performance to clinicians.

**Methods:** A cross-sectional study with qualitative analysis was conducted. Two clinical psychologists developed 22 psychiatric emergency vignettes. Responses were generated by three versions of GPT-4 (GPT-4o, GPT-4o Mini, GPT-4 Legacy) using ChatGPT, and two independent nurse practitioners (clinicians). The responses focused on three triage criteria: risk (Low 1-3 High), admission (Yes-1; No-2), and urgency (Low 1-3 High).

**Results:** Substantial interrater reliability was observed between clinicians and GPT-4 responses across the three triage criteria (Cohen’s Kappa: Admission = 0.77; Risk = 0.78; Urgency = 0.76). Among the GPT-4 models, Kappa values indicated moderate to substantial agreement (Fleiss’ Kappa: Admission = 0.69, Risk = 0.63, Urgency = 0.72). The mean scores for triage criteria responses between GPT-4 models and clinicians exhibited consistent patterns with minimal variability. Admission responses had a mean score of 1.73 (SD = 0.45), risk scores had a mean of 2.12 (SD= 0.83), and urgency scores averaged 2.27 (SD = 0.44).

**Conclusion:** This study suggests that GPT-4 models could be leveraged as a support tool in mental health telephone triage, particularly for psychiatric emergencies. While findings are promising, further research is required to confirm clinical relevance.

## INTRODUCTION

Young adults and adolescents are currently experiencing a mental health crisis.^1–3^ One of the cornerstones of this crisis is a heightened level of suicide risk due to various factors such as social isolation, hopelessness and depression^4,5^. As a result, suicide is the leading cause of death for 10-24 year olds in The United States and requires immediate attention.^6^ Youth presenting with psychiatric emergency conditions can include suicide, assaultive or violent behavior, and acute mental status changes such as psychosis, intoxication, and extreme anxiety^7^. For youth experiencing psychiatric emergencies, Emergency Departments (ED) are safe areas to visit since they are required to treat all patients without obligation. Approximately half a million children visit the ED every year with urgent psychiatric emergencies^8^, however, ED’s often lack the resources to identify and manage patients in psychiatric populations.

Since visiting the ED is not always the most optimal solution for patients with acute psychiatric conditions, prehospital triage helps patients speak with a clinician to determine their best course of action. Prehospital triage is a process in which patients are prioritized by the severity of their condition and their expected resource needs – which is tied to the urgency of their condition^9^.

Mental health prehospital triage is a form of prehospital triage that concentrates more on the risk of harm to self or others in addition to the urgency of the situation. In mental health triage, classifying the severity of psychiatric emergency patients by determining clinical characteristics quickly and accurately is the one of the most important first steps^10^. Mental health emergency hotlines such as the 988 Suicide and Crisis Lifeline act as prehospital triage centers and play a vital role in managing patients with acute psychiatric conditions^11^. Trained responders on these lines can provide a variety of services such as giving emotional support, providing suicide risk assessments, giving referrals to treatment and even transferring patients to emergency services^12^. Mental health telephone triage (MHTT) requires clinicians to be competent in many different areas such as resource and time management, working knowledge of psychopharmacology, knowledge of community-based services and referrals, therapeutic approaches and interventions, along with many others^13^. As a result, clinicians cannot always provide consistent and accurate triage responses which can prevent patients with psychiatric emergencies from receiving effective treatment in a timely manner. Fortunately, there are clinician support tools that can ease the burden on clinicians such as knowledge-based decisional algorithms and data driven artificial intelligence models^14^.

Artificial Intelligence has been evident in being able to assist clinicians in triaging acute emergency conditions in other specialties^15,16^. Similarly, Machine Learning (an application of AI) assisted prehospital triage models have the ability to triage more effectively than conventional models^17^. AI has also shown promise in being a support tool in online mental health care in the form of chatbots providing immediate mental health access as well as with helping clinicians create better therapeutic applications for psychiatric disorders^18,19^. Despite this, AI’s applications in mental health are still in its infancy^20^, and based on our knowledge of the current literature, there are limited studies observing the ability of artificial intelligence, specifically Large Language Models (LLMs), in assisting with the prehospital MHTT of psychiatric emergency conditions.

The primary aim of this study is to compare the ability of one of the prominent LLMs, GPT-4 by OpenAI, to triage young adult patients with psychiatric emergencies to that of clinicians. A secondary aim of the study is to determine which GPT-4 model, GPT-4o, GPT-4o Mini or GPT- 4 Legacy is the most effective in being able to triage patients.

## METHODS

In this study, we investigated the ability of GPT-4 Models via ChatGPT to triage patients with psychiatric emergencies based on risk, admission and urgency. To test different psychiatric emergencies, authors used ChatGPT to simulate a list of 22 clinical vignettes (see Appendix A) across five categories of common psychiatric emergencies: (1) psychosis, (2) suicidal ideation, (3) substance abuse, (4) extreme anxiety, (5) violent/destructive behavior^7^. Each vignette template had a paragraph format including information about age, race, gender, medication history, behavioral symptoms, and patient history (Figure 1).

**Fig.1.**
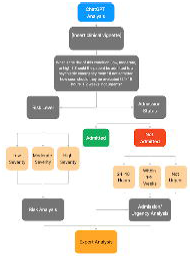

## Model Selection

GPT-4 was the LLM selected for this study due to its easy accessibility and its availability to the public. We decided to test the three different GPT-4 models: GPT-4o Mini, GPT-4o, and GPT-4 Legacy since they were the most up to date GPT models when this study was conducted^21^. We will refer to these three models as “GPT Models” from this point forward.

## Question Creation

The questions were formatted to represent a prehospital triage scenario in which the caregiver of the young adult would report the following information to a psychiatric crisis line or emergency department line. (Textbox 1) Each vignette was written in a paragraph format to reflect concerns about the individual’s behavior from the perspective of the caregiver of the young adult with a mental health condition from one of the five categories. From the original vignettes, vignette number 13 was disregarded due to disagreement between clinicians.

#### Textbox 1. Vignette Template

Tim is a 24 year-old PhD candidate at a large university. Over the past six months, his behavior has changed and become increasingly bizarre. Though originally very enthusiastic about graduate school, he states that he is no longer interested in pursuing a degree and has no motivation to continue with school. He used to drink alcohol socially, but has withdrawn his friends and is not currently using any substances. He has no history of drug or alcohol abuse. He is unable to concentrate on work and tells friends and family that he believes someone has been following him when he leaves the house, and spying on him in his bedroom at night.

Police bring him to a state psychiatric hospital after he throws his cell phone against a bus window and causes a public disturbance. Hospital staff members comment that they often hear him whispering frantically when he is alone, as though he is having a conversation with another person.

When asked about his behavior, Tim states the following:" I couldn’t. I couldn’t use my phone anymore - they’re listening… and I just couldn’t.""The agents are listening. They’re after me and I can’t get away.""I am wanted by the government. I’m sitting in an office building right now and I see tear gas coming up from the floor. Can you smell the poison they’re releasing into the air?" "The president wants to kill me, the agents all tell me it’s useless to fight it… I can’t go on.""I was born to end the world and bring down the government in this dismal country." "I had to stop drinking because my cups are all poisoned by government agents." The psychiatrist diagnoses Tim with schizophrenia and prescribes an antipsychotic. After 5 weeks on the medication, Tim shows minor improvements but the changes are not drastic. He continues to claim that he is a target of the government and is meant to bring down the country’s infrastructure. He occasionally claims to see agents hiding under the floorboards and above his head in ceiling tiles. In addition, he refuses to drink any beverages because he believes they have been poisoned. He is required to stay at the psychiatric hospital and receive treatment as an inpatient, because his psychiatrist believes he is a danger to himself and others^21^

### Data collection

Two psychologists and a psychiatrist on the research team reviewed the vignettes to create scenarios similar to what patients potentially experience during psychiatric crises. The vignettes along with the three questions seen in Figure 2 were entered into GPT-4o, GPT-4o mini and GPT-4 Legacy one time each and the subsequent responses were recorded for data collection into an Excel sheet for a total of 66 responses amongst the different GPT models (see Figure 1). The responses from all three models were recorded each time for expert evaluation and below is a sample response from GPT-4o (see Textbox 2). The same vignettes were then evaluated by two advanced practice nurses. Instead of following the same flowchart as the GPT models, the nurses were simply asked to rate each of the response criteria on a 3 point Likert-type scale. Three psychiatrists then evaluated the responses by both GPT and the nurses qualitatively using a coding framework (see Table 1).

**Fig.2.**
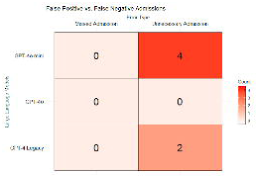

**Table 1.** Rubric for Content Analysis.

| Measures | Question | Description | Scale |
|---|---|---|---|
| Triage Accuracy | Is the answer an accurate reply to the question? | This criterion assesses the accuracy of the risk level, admission status and overall triage evaluation" **Accurate**" response fully answers the question with correct information. "**Inaccurate**" indicates the response is incorrect or irrelevant. "**Partially accurate**" suggests the response is on the right track but either contains some inaccuracies or doesn’t fully address all aspects of the question. | Accurate (3), Partially Inaccurate (2), Inaccurate |
| Clarity | Is the message clearly conveyed? | This measures how easily understandable the response is. "**Yes**" means the response is well-structured, easy to follow, and free of jargon or ambiguity. "**No**" implies the explanation is confusing, poorly structured, or uses overly complex language. "**Partially**" indicates the response has some clarity but may be improved in certain areas for better understanding. | Yes (3), Partially (2), No (1) |
| Completeness | Does the response completely answer the question? | This evaluates whether the response fully addresses all elements of the question. "**Complete**" indicates that the response covers all aspects of the question comprehensively. "**partial**" for partially complete missing some aspects. "**Incomplete**" means the response misses one or more critical elements of the question. | Complete (3), Partially Complete (2), Incomplete (1) |
| Overall Score | What is the overall score? | This rating reflects the confidence in the response's accuracy, clarity and completeness<br>**Low:** This rating suggests effectiveness and appropriateness of the response are likely significantly different from what is perceived. It indicates major issues in accuracy, clarity, inclusivity, completeness, or utility. Or this rating implies that there is a considerable possibility the response's actual effectiveness and appropriateness might differ significantly from what is perceived. While not entirely unreliable, the response has notable deficiencies.<br>**Moderate:** This rating is used when it is believed that the response is probably close to what is perceived in terms of accuracy, clarity, inclusivity, completeness, and utility. The response is reliable but might have minor areas for improvement.<br>**High:** This rating indicates a strong level of confidence in the response. It suggests that the true effectiveness and appropriateness of the response are very similar to what is perceived, showing high levels of accuracy, clarity, inclusiveness, completeness, and utility. | High (3), Moderate (2), Low (1) |

##### Textbox 2. Example generated response by ChatGPT-4o

**Vignette:** Emily is a 17-year-old Hispanic female high school senior who has recently been experiencing visual and auditory hallucinations. She reports seeing shadows and hearing whispers, particularly when she is alone or trying to fall asleep. Despite these troubling symptoms, Emily continues to perform well academically and is actively involved in her school’s drama club. Her parents have observed increased anxiety and some social withdrawal, but Emily has not exhibited aggression or experienced any academic impairment. She denies any thoughts of self-harm or harm to others. Emily has a history of occasional anxiety and was briefly on sertraline, which she stopped due to mild side effects. There is also a family history of anxiety and mild depression.

**Response**: *Emily’s risk level can be considered **moderate**. Although she is experiencing hallucinations, she continues to function well academically and socially, and she denies any thoughts of self-harm or harm to others. Her symptoms, though concerning, do not currently suggest an immediate risk of danger to herself or others, which reduces the urgency for psychiatric hospitalization*.

*Given the presence of hallucinations, increased anxiety, and social withdrawal, it is important that Emily be evaluated by a mental health professional in a timely manner. However, since there is no immediate threat or severe impairment, it is not necessary for her to be admitted to a psychiatric emergency room at this time*.

*The recommendation would be to have Emily evaluated within **1-2 weeks** by a psychiatrist or psychologist to further assess her condition and to explore potential interventions or treatment options. If her symptoms worsen or if there is any indication of self-harm, harm to others, or significant functional decline, an emergency evaluation should be sought sooner*.

Interaction pathway for GPT models (blue box). After inserting the clinical vignette (gray box one), GPT models were presented with a series of triage-related questions (gray box two) and responses were recorded and evaluated. The first criterion (Risk Level, gray box) classified responses into one of three different possible responses of risk scores (peach boxes: Low Severity, Moderate Severity, or High Severity). The second criteria (Admission Status, green or red box) evaluated whether the patient should be admitted to an emergency room (Admitted, green box) or not (Not Admitted, red box). For cases where GPT models indicated no direct admission (red box), responses were further categorized based on evaluation urgency (peach boxes: 24-48 Hours, Within 1-2 Weeks, or Not Urgent). Expert analysis (orange box) was then conducted to assess risk level and admission/urgency evaluation based on the responses.

### Evaluation Criteria and Measures

The evaluation criteria we created in this study was used to assess the performance of each of the GPT models and clinicians. The criteria was created based on literature examining the clinical relevance of LLM tools^23,24^. The Expert Evaluation Criteria included Triage Accuracy, Clarity, Completeness, and Overall Score (see Table 1). Experts also provided feedback in the form of text responses on the different responses by the GPT models as well as on their ratings. This feedback was recorded in the same Excel sheet used to record the responses by GPT models.

The authors used a systematic approach to grade each response. All responses were graded on the five criteria on the 3-point scale, with 3 representing the best fulfillment of that specific criterion. For example, a three on triage accuracy would indicate that the GPT model was able to correctly triage a certain patient fully based on risk level, admission status and urgency of evaluation (see Figure 1). Conversely, a one on completeness would indicate that the response by the GPT model was incomplete and did not address the requirements of the question.

### Data preparation

Preprocessing of study data included reverse scoring Urgency (originally High 1-3 Low) to match the scale of Risk (now Low 1-3 High) for interpretability. Partial or range (i.e., “1-2” or “2-3”) answers were interpreted as an average (i.e., 1.5 or 2.5 respectively) then rounded up. Additionally, Urgency scores that were originally “0” based on the dependence of Admission being “1” were interpreted as “NA” and were not calculated during analysis.

### Analysis selection

We assessed agreement of clinicians and large language models using two methodologies. First, we separated them into 2 groups to determine the Cohen’s Kappa (weighted quadratically). Since healthcare providers can vary slightly in their approach to admissions, risk assessment, and urgency by discipline, we used quadratic weights to penalize extreme differences more harshly between clinicians and GPTs. Second, we compared the 3 models within the GPT group for agreement with Fleiss’s Kappa. Descriptive analysis was conducted by splitting the overall dataset into 3 response types.

In terms of the Kappa coefficients, ≤ 0 indicated no agreement, 0.01 to 0.20 indicated slight ≤ agreement, 0.21-0.40 indicated fair agreement, 0.41-0.60 indicated moderate agreement, 0.61-0.80 indicated substantial agreement, and 0.81-1.00 was indicative of nearly perfect agreement^25^.

The Cohen’s Kappa statistic was used for two raters and the Fleiss Kappa statistic was used for more than two raters in the case of comparing the three different GPT models.

### RESULTS

The GPT models overall had strong responses to the clinical vignettes overall had a strong level of triage accuracy, clarity, completeness and overall score. Findings are presented using the 3- point scale evaluated by the psychiatrists (see Table 2). In this study, the different GPTmodels were also compared against each other. Regarding Triage Accuracy, GPT-4o had the highest mean score (M = 2.56, SD = 0.68) which was equivalent to GPT-4 Legacy (M = 2.39, SD = 0.74), and GPT-4o mini had the lowest mean (M = 2.39, SD = 0.65). Following this, Clarity scores were the highest for GPT-4o (M = 2.65, SD = 0.48) which was also equivalent to GPT-4 Legacy (M = 2.65, SD = 0.48) and the lowest was GPT-4o mini (M = 2.36, SD = 0.54).

**Table 2.** LLM tools organized by Expert Evaluation Criteria.

| **Model** | **Triage Accuracy** | **Clarity** | **Completeness** | **Overall** |
|---|---|---|---|---|
| **GPT-4o** |  |  |  |  |
| M | 2.56 | 2.65 | 2.45 | 2.53 |
| SD | 0.68 | 0.48 | 0.56 | 0.61 |
| Mdn | 3.00 | 3.00 | 2.00 | 3.00 |
| IQR | 1.00 | 1.00 | 1.00 | 1.00 |
| **GPT-4o Mini** |  |  |  |  |
| M | 2.39 | 2.36 | 2.20 | 2.29 |
| SD | 0.65 | 0.54 | 0.66 | 0.70 |
| Mdn | 2.00 | 2.00 | 2.00 | 2.00 |
| IQR | 1.00 | 1.00 | 1.00 | 1.00 |
| **GPT-4 Legacy** |  |  |  |  |
| M | 2.56 | 2.65 | 2.48 | 2.47 |
| SD | 0.74 | 0.48 | 0.50 | 0.68 |
| Mdn | 3.00 | 3.00 | 2.00 | 3.00 |
| IQR | 1.00 | 1.00 | 1.00 | 1.00 |

Min = 1, Max = 3

Completeness scores were the highest for GPT-4 Legacy (M = 2.48, SD = 0.50) followed by GPT-4o (M = 2.45, SD = 0.56) and lowest for GPT-4o Mini (M = 2.20, SD = 0.66). Overall Score was the highest for GPT-4o (M = 2.53, SD = 0.61) followed by GPT-4 Legacy (M = 2.47, SD = 0.68) and again the lowest for GPT-4o Mini (M = 2.29, SD = 0.70).

As given in Table 3, The interrater reliability between clinicians and GPT models was substantial (Cohen’s Kappa: Admission=0.77 (95% CI, 0.70-0.84), Risk=0.78 (95% CI, 0.72-0.85), Urgency=0.76 (95% CI, 0.70-0.83), p<0.001). Among the GPT models, Kappa values also reflected moderate to substantial agreement (Fleiss’ Kappa: Admission=0.69, Risk=0.63, Urgency=0.72). These Kappa values suggest a high level of concordance, showing substantial agreement across both clinicians and GPT models. Descriptive analysis showed that the mean scores were 1.73 (SD = 0.45) for Admission, 2.12 (SD = 0.83) for Risk, and 2.27 (SD = 0.44) for Urgency, with minimal variability and consistent scoring patterns across all categories.

**Table 3.** Interrater reliability score between clinicians and GPTs , among GPT models and descriptive statistics.

| Triage Measures | Clinicians & GPTs (Cohen’s Kappa W<sup>2</sup>) | 4o, Mini, Legacy (Fleiss) | 95% CI (Cohen’s Kappa W<sup>2</sup>) | Mean (SD) | Min-Max | Range |
|---|---|---|---|---|---|---|
| Admission | 0.77* | 0.69* | 0.70-0.84 | 1.73 (0.45) | 1-2 | Yes 1-2 No |
| Risk | 0.78* | 0.63* | 0.72-0.85 | 2.12 (0.83) | 1-3 | Low 1-3 High |
| Urgency | 0.76* | 0.72* | 0.70-0.83 | 2.27 (0.44) | 1-3 | Low 1-3 High |

\*p<0.001

This heat map displays the distribution of errors for admission decisions across the three GPT models (GPT-4o, GPT-4 mini and GPT-4 Legacy) categorized into two error types (Missed Admission and Unnecessary Admission. The color intensity represents the count of the error, with darker shades of red indicating higher count values and lighter shades of red indicating lower counts. The lowest count of “0” represents no errors for that category. The color scale ranges from (count = 0) to (count = 4) as shown in the accompanying color bar.

In our study, false positives occurred when LLMs admitted patients when the clinicians did not admit patients resulting in unnecessary admission. On the other hand, false negatives were when clinicians admitted patients when the LLMs did not which results in a missed triage patient. We found that there were more total false positives compared to false negatives across LLMs. In fact, there were no false negatives at all (see Figure 2). The LLMs in overall responses leaned towards admitting more patients compared to not admitting at all.

### DISCUSSION

This study evaluated the performance of three GPT models in generating responses to clinical vignettes based on five common categories of psychiatric emergencies experienced by young adults and compared the performance to that of two nurse practitioners. We observed substantial agreement between clinicians and GPT models as well as significant differences between GPT- 4o mini and the other GPT models in the following metrics: Triage Accuracy, Clarity, Completeness, and Overall Score.

In terms of Triage Accuracy, GPT-4o and GPT-4 Legacy were both shown to outperform GPT- 4o mini. Previous studies have observed similar results where GPT-4o Mini may compromise accuracy and consistency in responses when answering multiple-choice exams in different medical specialties^26,27^. Clarity and completeness were also areas where GPT-4o Mini underperformed in comparison with GPT-4o and GPT-4 Legacy which also has been observed in additional research evaluating the clinical capabilities of both models in managing lumbar disc herniations^28^. This can be attributed to the fact that it is an LLM tool meant for efficiency and speed for greater accessibility as opposed to consistency in response^29^. As a result, it is no surprise that GPT-4o Mini had the lowest overall score out of the three GPT models. While the speed of information retrieval is crucial during crisis calls, these situations often involve sensitive and complex issues that demand clear and thorough responses. By collecting and organizing high-quality triage information, GPT models can support clinicians in making well informed decisions to improve the overall effectiveness of telemental health triage systems.

Interrater reliability (IRR) between clinicians and GPT models was observed with moderate to substantial agreement in admission (κ = 0.77), risk (κ = 0.78), and urgency (κ= 0.76). This IRR indicates that GPT models can replicate much of the reasoning that clinicians use in decision making and can support clinical workflows by offering consistent and reliable assessments for the majority of cases, but discrepancies may arise in edge cases. Since LLMs use probabilistic reasoning as opposed to human intuition there are certain contextual factors that contribute to variability in clinician-decision making that LLMs are limited in recognizing^30^. Suicide risk assessment is an example of a tool that relies on understanding contextual factors such as comorbid conditions and the interplay between mental health and social determinants of health - areas that may exceed the scope of understanding that LLMs can comprehend. Calculated risk scores from suicide risk assessment tools have a low predictive ability that represents a challenging gray area for GPT models since they requires nuanced judgement beyond direct calculation and merely serve as an aid for clinician-decision making^31^.

Compared to clinicians, we noticed that GPT models on average admitted more patients than necessary. GPT models’ tendency to over admit patients may stem from the model’s conservative design, which prioritizes minimizing the risk of adverse outcomes in triage^32^. This was seen mostly from GPT-4o Mini which had four unnecessary admissions and GPT-4 Legacy that had two unnecessary admissions. Interestingly, we noticed that GPT-4o did not overadmit any patients and we believe this could be due to the fact that the model had been tested by experts on social psychology, bias and fairness and misinformation introduced by new modalities^33^. However, compared to clinicians, GPT models overall appear to overvalue the possibility of less severe mental health crises and recommend immediate referral to the ED, even in cases where less severe presentations could be appropriately managed through outpatient follow-up or scheduled evaluations over longer periods of time^34^. The inability to accurately assess levels of risk makes it difficult to consider alternative options. Referrals to community health programs, therapy or outpatient settings can offer valuable support but without thorough assessment, these options are overlooked, creating additional strain for EDs^35^.

Our findings indicate that GPT models can serve as a triage support tool for clinicians by evaluating patients with psychiatric emergencies. Given the ongoing youth mental health crisis and the shortage of behavioral health care professionals, there is an increasing need for technology-supported screening methods that enhance efficiency and accessibility^36^. Computer assisted screening tools already exist in screening for psychiatric disorders in mental health telephone triage, but don’t take into account real-time interactions^37^. Real-time feedback is crucial in mental health prehospital triage since it is a time-sensitive process and mental health emergencies require timely and effective care. By being able to access real-time analysis of a patient’s condition, clinicians can implement faster deliveries of interventions to prevent the escalation of psychiatric emergencies.

However, such AI tools have their own limitations. It cannot have the “human touch” that clinicians can provide, but can be complementary support which can allow clinicians to focus on delivering empathetic care^38^. Clinicians require many different competencies during MHTT, but building rapport and communicating effectively with patients are key core competencies of MHTT and overall telephone triage that only a human can provide^13^. This process can often be difficult and time-consuming for clinicians coupled with the effect that adolescents are apprehensive to approach mental health services, and have generally negative attitudes toward mental health professionals^39^. Having an AI assistant can help clinicians conduct risk assessments and perform mental health status examinations which are core competencies of MHTT that do not involve directly interacting with the patient. GPT models are not capable of providing accurate and consistent information on their own yet since they frequently “hallucinate” and make reasoning errors^40^, but they have the potential to help guide clinical examination and compliment clinicians in the future. The integration of clinicians and artificial intelligence in this way has the potential to significantly enhance the overall MHTT process by combining the strengths of human expertise with the efficiency and support provided by GPT models.

### Limitations and Future Considerations

There are several limitations to this study. First, clinical vignettes can potentially serve as a starting point to identifying GPT models as a potential triage support tool, but further research will need to incorporate real patient data. Our study focused on a limited range of psychiatric emergency categories and utilized controlled scenarios that do not fully capture the complexity and comorbidity often presented in real-life patient cases. MHTT is also a relatively novel concept and it is not a standardized protocol across emergency hotlines or crisis lines.

Additionally, the evaluation criteria used in this study was extended from previous studies involving the assessment of LLMs and has not been validated. The moderate IRR between experts indicated that there was potentially a difference in interpretation of the criteria, and experts also did not evaluate clinician responses to the clinical vignettes since the components of the triage accuracy criterion (severity, admission, and urgency) is largely subjective.

Clinical vignettes are not representative of an interaction between a clinician and a patient which is a lot more nuanced and has more variability. The flowchart we used is an extremely simplified version of how patient data could be assessed by GPT models during a call. The capacity of GPT models to provide real-time feedback was also not evaluated since patient information was presented all at once. Our study was also limited to only one LLM although we did compare and contrast the capability of three different versions: GPT-4o, GPT 4o Mini and GPT-4 Legacy.

GPT models and other LLMs are constantly evolving and as a result the performance of future LLM models could indicate different findings over time. Future considerations could be using other LLMs such as Cohere, Microsoft Copilot and Gemini as well as using up to date GPT models (GPT-o1, GPT-o1 Mini).

### Conclusion

Our study contributes to the growing body of literature of AI applications in healthcare by addressing how GPT models can evaluate patients with acute psychiatric conditions or emergencies. By comparing the performance of these models to that of clinicians we were able to evaluate the ability of AI to work as a prehospital triage support tool. The findings of this study provide valuable insights for future research on the application of GPT models in mental health telephone triage and contribute to advancements in AI for mental health care.

### Supporting information

Supplemental Table 1

## Data Availability

All data produced in the present study are available upon reasonable request to authors

## ACKNOWLEDGEMENTS

No funding declared. K.H., K.L. and M.Y. composed of the expert committee and analysed the GPT models’ responses. E.S. (Erin Sunderland) and S.S. were the two nurse practitioners in the study and helped analyze the clinical vignettes.

## DISCLOSURE STATEMENT

None declared

## AUTHOR CONTRIBUTIONS

S.T. conceived the project, collected data, and wrote the manuscript. M.Y. contributed to project conception, assisted with data collection, and organized the study. E.S. and D.J. analyzed the data and designed the analyses. I.M. and W.L. reviewed the clinical vignettes and contributed to the manuscript. E.Y. supervised the project and provided guidance on the analyses and overall manuscript.

### Submission Field

This manuscript is submitted under the primary field general psychiatry and related fields because it covers the topic of psychiatric emergencies and the applications of artificial intelligence in psychiatry. The secondary field that this manuscript is being submitted is under child and adolescent psychiatry is infant, child, and adolescent psychiatry since the clinical vignettes involve adolescents and young adults.

Our manuscript is not a clinical trial paper because we used clinical vignettes which are case studies designed to simulate real life patient scenarios.

## Notes

### Competing Interest Statement

The authors have declared no competing interest.

### Funding Statement

This study did not receive any funding

### Author Declarations

This study used simulated data that were openly available to the public before the initiation of the study

