## Supplemental Table 1 for "Potential of ChatGPT in Youth Mental Health Emergency Triage: Comparative Analysis with Clinicians"

Appendix A.

| Category | Vignette Description |
| --- | --- |
| Psychosis | Emily is a 17-year-old Hispanic female high school senior who has recently been experiencing visual and auditory hallucinations. She reports seeing shadows and hearing whispers, particularly when she is alone or trying to fall asleep. Despite these troubling symptoms, Emily continues to perform well academically and is actively involved in her school's drama club. Her parents have observed increased anxiety and some social withdrawal, but Emily has not exhibited aggression or experienced any academic impairment. She denies any thoughts of self-harm or harm to others. Emily has a history of occasional anxiety and was briefly on sertraline, which she stopped due to mild side effects. There is also a family history of anxiety and mild depression. |
| Psychosis | Jordan is a 19-year-old Black male college freshman who has experienced significant mental health changes over the past three months. He reports frequent auditory hallucinations, such as hearing voices, and has developed paranoid thoughts, believing that others are plotting against him and monitoring his actions. These symptoms have led to social withdrawal, neglect of personal hygiene, and a marked decline in academic performance. Although Jordan denies thoughts of self-harm or harm to others, his symptoms are causing noticeable distress and functional impairment. His medical history includes a prior episode of depression managed with fluoxetine, which was discontinued due to side effects, and there is no history of substance abuse. |
| Psychosis | Alex is a 22-year-old Asian American male who has been exhibiting psychiatric symptoms over the past two months. He reports experiencing intense auditory and visual hallucinations, including hearing commanding hallucinations that tell him to harm himself and seeing frightening, distorted images. Alex has also developed significant paranoia, believing he is being followed and that his phone is being monitored. His behavior has become erratic, including instances of aggression towards himself and others, and he has withdrawn completely from daily responsibilities and social interactions. His medical history includes a previous diagnosis of bipolar disorder, for which he was prescribed lithium and quetiapine but has been non-compliant with medication for the past six months. There is a family history of schizophrenia and severe mood disorders. |
| Psychosis | Lena is a 20-year-old White female college student who has been experiencing psychiatric symptoms over the past three months. She reports hearing faint voices that occasionally criticize her and feeling a persistent sense that people are watching her, especially in public places. These symptoms have caused her to become more socially withdrawn, avoiding gatherings with friends and skipping some classes. Despite these challenges, Lena continues to maintain her grades, though with increased difficulty and stress. Her medical history includes generalized anxiety disorder, for which she was prescribed escitalopram, but she stopped taking it a few months ago on her own. There is no history of substance abuse, but there is a family history of depression and anxiety. |
| Suicidal Ideation | Maya is a 17-year-old Latina high school junior who has recently been experiencing episodes of sadness and stress related to school and social pressures. Over the past few weeks, she has had occasional fleeting thoughts of whether life is worth living, particularly during moments of feeling overwhelmed, but she immediately dismisses these thoughts and does not have any plans or intentions to harm herself. Maya continues to perform well academically and remains active in her school’s soccer team and volunteer activities. Her parents have noticed that she has been more reserved than usual but is still engaging in family activities. Maya’s medical history is unremarkable, with no prior psychiatric diagnoses or history of substance use. There is a family history of mild depression on her mother’s side. |
| Suicidal Ideation | Sam is a 19-year-old Black male college sophomore who has been struggling with increasing feelings of hopelessness and sadness over the past two months. He reports having recurrent thoughts of suicide, particularly during periods of intense stress or after experiencing personal setbacks, such as a recent breakup and academic difficulties. While Sam has thought about specific methods, he has not made any plans to act on these thoughts. He often isolates himself in his dorm room, neglecting schoolwork and avoiding social activities, though he still attends some classes. Sam's friends have noticed that he has been withdrawing and expressing more negative thoughts about his future. His medical history includes a previous episode of depression during high school, for which he was briefly treated with cognitive-behavioral therapy. There is a family history of depression and substance use. |
| Suicidal Ideation | Aisha is a 21-year-old African American female college senior who has been experiencing depressive symptoms over the past three months. She reports having frequent and intense thoughts of suicide, including specific plans to overdose on medication. Aisha has begun writing goodbye letters to her family and friends and has recently withdrawn from almost all activities, including her classes and social life. Her sleep has become increasingly disturbed, and she often stays in bed for most of the day. Aisha’s friends have noticed drastic changes in her behavior, including expressing feelings of worthlessness and hopelessness. Her medical history includes a diagnosis of major depressive disorder, for which she was prescribed sertraline, but she stopped taking it several months ago without consulting her doctor. There is a family history of depression and suicide on her father's side. Aisha is covered by her parent's health insurance plan through a PPO. |
| Suicidal Ideation | Kevin is a 23-year-old Hispanic male college graduate who recently expressed dramatic statements about life not being worth living during a heated argument with his roommate. These statements alarmed his friends and family, prompting concerns about his mental well-being. However, upon further conversation, Kevin clarified that he does not have any actual intent to harm himself nor does he have a plan to do so. He mentioned that his remarks were more about expressing frustration with his current unemployment and recent breakup than a true desire to end his life. Kevin remains active in his social circles and continues to engage in hobbies like basketball and graphic design, which he finds uplifting. He has no history of mental health issues and no family history of suicide. Kevin is covered by health insurance through his last employer, which is still active. He has agreed to see a counselor to discuss better ways to handle stress and emotional expression, recognizing the impact of his words. |
| Suicidal Ideation | Lily is a 24-year-old Asian female graduate student who has been experiencing persistent feelings of sadness and occasional thoughts of self-harm over the past six months. While Lily has not formulated any plans for self-harm, she often feels overwhelmed by thoughts of not being good enough and worries she may eventually feel compelled to act on these thoughts. Her academic performance has declined, and she finds it difficult to concentrate on her studies or engage in conversations with her peers. Lily frequently misses days at university, citing lack of motivation and energy as her main reasons. She has a history of anxiety diagnosed during her undergraduate years, for which she was prescribed escitalopram, but she took it intermittently and has not sought any treatment recently. Lily has expressed to a close friend that she feels stuck and unsure how to change her situation. She has a family history of depression. Lily is covered under a student health insurance plan. |
| Suicidal Ideation | Jessica is a 22-year-old White female who has recently completed her Master's degree in Public Health. To those around her, Jessica seems to be thriving: she has started a competitive internship, remains socially active, and frequently attends yoga and meditation classes. Despite these achievements, Jessica is grappling with persistent thoughts of suicide. She has meticulously planned various methods and times for potentially ending her life, without sharing these thoughts or her growing sense of despair with anyone. Her ability to maintain a composed and engaged demeanor has effectively concealed her struggles. Jessica uses her extensive knowledge of mental health to mask her symptoms, ensuring that her friends and family see only her successes and none of her internal turmoil. Jessica is covered by health insurance through her recent graduate program, but she has not yet sought professional help for her mental health issues, believing she should be able to manage on her own given her background in public health. |
| Substance Use | Tara is a 20-year-old Caucasian female college sophomore who has recently started experimenting with cannabis. She uses it primarily on weekends when socializing with friends and denies using it alone or feeling a need to use it to function daily. Tara maintains a good academic record and is active in several clubs, including the university debate team. Her use has not led to any negative consequences in her studies or personal life, and she remains highly engaged in her academic and social activities. Tara has no prior history of substance abuse, and there is no significant family history of substance use disorders. She is aware of the potential risks associated with regular use and expresses a commitment to keeping her use recreational and controlled. Tara is covered by a private health insurance plan through her parents. |
| Substance Use | Nathan is a 21-year-old Asian American male college senior who recently became the focus of concern among his dorm's resident advisor and campus security after being found with a considerable amount of marijuana in his dorm room. Initially suspected of selling marijuana, Nathan explained that his regular use is primarily for stress relief during exams and is shared only with close friends. Despite the seeming severity of the situation, Nathan's academic performance remains stable, and he holds a leadership position in his robotics club. His use, while more frequent than ideal, has not led to serious personal or academic consequences. Nathan does not exhibit any patterns of deceit, aggression, or rule-breaking beyond this incident, and his marijuana use appears to be isolated to personal stress management rather than indicative of antisocial or conduct disorder. He is covered by his university’s health insurance, which includes mental health and substance abuse services, and has decided to seek counseling to address his stress and reassess his use of marijuana, aiming to ensure it does not impact his final year of studies. |
| Substance Use | Derek is a 20-year-old Black male college sophomore whose drug use has escalated to include cocaine and opioids. His behavior has become increasingly problematic, marked by secrecy, financial instability, and a sharp decline in academic performance. Derek insists he has control over his drug use, yet his actions suggest a deepening dependency that affects every aspect of his life. His situation is complicated by a family history of substance abuse, increasing the urgency for intervention. He has resisted seeking help, raising concerns among friends and faculty about his well-being and future. |
| Substance Use | Ella is a 19-year-old White female college freshman who was recently caught smoking marijuana in her dorm room. The incident, combined with her frequent late-night social activities, led to widespread rumors among her peers and some faculty members about a potential substance abuse problem. Concerned, her resident advisor reported the situation, which prompted a meeting with college administrators. During the meeting, Ella explained that she occasionally uses marijuana socially, primarily at gatherings with friends, and does not use it alone or feel a compulsion to use it. Her academic performance remains strong, and she actively participates in extracurricular activities, including the debate team and a volunteer group. Despite the initial concerns, further discussions revealed that Ella’s use was infrequent and not interfering with her responsibilities. She has no history of substance abuse, and her health is generally good. Ella’s family has no significant history of substance abuse disorders. She is covered by her parents' health insurance plan, which includes mental health services. |
| Extreme Anxiety | Ben is a 21-year-old Caucasian male university student who recently visited the emergency room during a final exam due to what he described as the "worst panic attack of [his] life," experiencing severe chest pain, shortness of breath, and a feeling of impending doom. Although the episode appeared severe and prompted an immediate medical response, medical professionals determined that Ben's symptoms were purely anxiety-related with no underlying cardiac issues. These panic attacks, while intense, were infrequent and occurred primarily in high-stress situations like exams. Outside these instances, Ben functions well, maintaining strong academic performance and active involvement in sports and social activities. He has no history of chronic anxiety or other mental health disorders, and his physical health is excellent. Ben is covered by his university's student health insurance, which includes mental health services. |
| Extreme Anxiety | Liam is a 20-year-old Caucasian male college junior experiencing severe anxiety that has increasingly disrupted his life over the past year. His symptoms include frequent panic attacks, an intense fear of public speaking, and persistent worries that significantly impair his daily activities and sleep. Unable to attend classes regularly due to fear of interaction in large groups, Liam's academic performance has sharply declined. He avoids social situations and spends most of his time isolated in his dorm room. Despite attempting to manage his anxiety through online resources and breathing exercises, these methods have provided minimal relief. With no prior history of mental health issues, the severe onset of symptoms has alarmed his friends and family. Covered by his parents' health insurance, which includes mental health services, Liam is now recognizing the necessity of seeking professional help as his self-management efforts are insufficient. |
| Extreme Anxiety | Mia is a 19-year-old Asian American female college sophomore who has been experiencing severe anxiety. Her condition has escalated to daily panic attacks and debilitating symptoms such as shaking, rapid heartbeat, and hyperventilation, significantly impairing her ability to attend classes or socialize. Mia's academic performance has plummeted, and she is unable to participate in group activities or public gatherings for fear of a panic attack. Attempts at managing her anxiety through relaxation techniques and online therapy have proven ineffective. Previously experiencing only mild anxiety, her current state is a stark escalation. Mia’s family, deeply concerned, is supportive and actively seeking enhanced mental health treatments. She is covered under her university's comprehensive health insurance plan, which includes mental health services, potentially facilitating access to more specialized care. |
| Violent/Destructive Behavior | Tyler is a 17-year-old Caucasian male high school junior who recently alarmed his teachers and parents with an outburst in class where he knocked over a chair and yelled at a peer. This incident, which seemed severe due to its public nature, led to a swift intervention by school officials. Despite this outburst, Tyler's behavior is typically non-violent and his academic performance remains solid. Discussions with school counselors revealed that Tyler's actions were an isolated response to extreme frustration from personal issues at home, rather than an indication of a chronic behavioral problem. He has no history of recurrent aggression, and his overall demeanor at school is cooperative and engaged. His parents are arranging for him to see a therapist to address his stress management, supported by their health insurance, which includes coverage for mental health services. Tyler and his family are committed to understanding and improving his responses to stress, reinforcing the incident as an outlier rather than a regular occurrence. |
| Violent/Destructive Behavior | Cara is a 16-year-old African American female high school sophomore who has exhibited occasional violent behavior, such as throwing objects and shoving classmates, which has caused concern among her teachers and peers. Cara has never used any substances in the past. These episodes occur a few times a month and seem to be triggered by interpersonal conflicts or high stress situations at school. Despite these incidents, Cara maintains average grades and participates in the school’s basketball team. She has expressed remorse following each episode and recognizes the need for better emotional regulation. Discussions with a school counselor indicate that Cara may be struggling with unresolved anger and stress management issues, rather than a more pervasive conduct disorder. Her parents are supportive and have involved her in weekly therapy sessions covered by their health insurance, focusing on developing coping strategies and emotional expression skills. Cara’s situation is being closely monitored to ensure her behavior does not escalate and to support her positive involvement in school activities. |
| Violent/Destructive Behavior | Lucas is an 18-year-old Hispanic male high school senior whose mild aggressive behaviors, such as snapping at classmates and being dismissive towards teachers, have become publicly noticed at school. Lucas also harbors intense feelings of anger and frustration, leading him to vandalize school property late at night. Although these incidents are known to some of his peers and faculty, his academic performance has only slightly declined, which hasn’t alarmed his teachers significantly. Publicly, Lucas attributes his outbursts to bad days or just reactions to stress. However, a close friend has publicly expressed concern over Lucas's increasing detachment and mentions of feeling overwhelmed, pointing to deeper emotional turmoil. Lucas’s family and the school community view these incidents as isolated and not indicative of a larger problem. He is covered under his family's health insurance, which includes mental health services, yet Lucas has been resistant to seeking help, maintaining that he can manage on his own. |
| Violent/Destructive Behavior | Emma is a 16-year-old Caucasian female high school sophomore who has been showing aggressive behaviors, such as shouting at teachers and pushing a classmate during an argument, multiple times over the past month. These outbursts have caught the attention of school administrators and her peers. Although Emma continues to participate in school activities and maintains average grades, her teachers have noticed a decline in her enthusiasm and a reluctance to engage in group work. The behavior appears linked to increased stress at home and anxiety about school performance, which she occasionally discusses with friends. Aware of the issues, her parents have been contacted by the school and are actively looking to connect her with a therapist under their health insurance plan, which includes mental health services, to help Emma develop better coping mechanisms and improve her behavioral responses. |
| Violent/Destructive Behavior | Nolan is a 17-year-old Black male high school junior who has recently been reprimanded for disruptions in class, such as talking out of turn and occasional inattentiveness. These behaviors, while noticeable, have not significantly impacted his overall academic performance or social interactions at school. Nolan maintains a B average in his classes and is a contributing member of the school's track team. The incidents are often attributed to lapses in concentration rather than any intent to disrupt. Nolan's teachers and his coach have discussed these behaviors with him, emphasizing the importance of staying focused, but they recognize these as typical teenage behaviors. Nolan's family is supportive, and he has a strong network of friends. He is covered under his parents' health insurance plan, which includes mental health services, though there has been no need to utilize these services as his behaviors are currently viewed as manageable. |
